# NICU Voices: Understanding Parent Perspectives of Research in the Neonatal Intensive Care Unit

**DOI:** 10.1101/2025.03.12.25322895

**Authors:** Melissa Coloma, Brandon Nguyen, William Cody Bartrug, Louie M. Swander, Karla Luna Silva, Egbert Villegas, Adam Numis, Patrick McQuillen, Shabnam Peyvandi, Elizabeth E. Rogers, Elizabeth E. Crouch, Mercedes Paredes

## Abstract

**Importance:** Neonatal intensive care units (NICUs) care for a vulnerable population with suboptimal research recruitment rates. Understanding NICU parents’ motivations and recommendations may improve recruitment efforts.

**Objective:** Identify key factors influencing NICU parents’ decisions to enroll their newborns in research and gather recommendations to enhance engagement.

**Design:** Semi-structured interviews were conducted with 24 parents from three NICU study populations: NSR-RISE, TRANSIT-CHD, and PROMPT. Using a grounded theory approach, data was analyzed prior to developing hypotheses, allowing themes to emerge organically during data analysis. Transcripts were coded through multiple rounds of data analysis until thematic saturation was reached.

**Setting:** Interviews occurred virtually with previous research participants at UCSF hospitals.

**Participants:** 65 parents of NICU patients were invited; 24 participated. Inclusion criteria included 1) parent age older than 18 years, 2) NICU admission history, 3) prior participation in NSR-RISE, TRANSIT-CHD, or PROMPT, and 4) child aged 18-36 months at time of interview.

**Main Outcome(s) and Measure(s):** Using a grounded theory approach, data was analyzed prior to developing hypotheses, allowing themes to emerge organically during data analysis.

**Results:** Parents of 8 NSR-RISE, 8 TRANSIT-CHD, and 8 PROMPT-enrolled neonates participated. Three primary themes emerged: 1) parents’ lived experiences during an emotionally intense NICU period fostered parental resilience and newfound support systems, 2) decision-making regarding NICU research participation included factors such as prognosis, emotional state, desire to aid future families, and perceived risks versus benefits, and 3) recommendations for improving NICU research recruitment, such as timely, empathic communication from trusted researchers, sensitivity to emotions, concise language, and early emphasis of altruistic goals.

**Conclusions and Relevance:** Altruism is a key motivator for NICU parents’ research participation. Recruitment strategies should emphasize empathetic, well-timed communication from trusted persons, clearly addressing risks and altruistic outcomes. Sensitivity to the emotionally charged NICU environment is essential for improving engagement and enhancing the NICU experience.

## Introduction

Participation in children’s health research may have beneficial effects across entire family units. Pediatric research has unique challenges, including ethical and logistical barriers that complicate obtaining consent from guardians, ultimately contributing to reduced research participation.^1^ The limited involvement of pediatric patients, particularly neonates, in research has far-reaching consequences for evidenced-based care, leaving significant gaps in understanding best practices for treating complex conditions in newborns. However, when presented with hypothetical situations, a majority of caregivers expressed willingness to enroll their child in research^3^, particularly when recommended by a trusted physician.^1,4^ Even in the profound setting of neonatal loss, some parents desire to contribute to research because it allows them to engage in parenting their child and provides meaning.^13^

Recruitment rates in neonatal populations remain suboptimal, leading to limited research engagement. This perpetuates a cycle of decreased understanding of the research process, reduced potential for generalization of clinical outcomes, and further hinders participation rates.^5,6^ Identifying the factors that influence parent’s decisions to enroll their children in research is critical for guiding future outreach efforts. Additionally, understanding the nuances of parental decision-making processes can provide researchers with actionable insights into timing and communication strategies during the recruitment process. Dahan et al. observed that parents of NICU patients primarily declined research invitations due to misunderstandings during the consent process.^2^ However, factors influencing a NICU parent’s enrollment may predate the consent process, potentially stemming from prior experiences, historical mistrust of the medical system, or cultural or language barriers. ^14^

There remains a need for further research into the NICU parent perspective across different stages of research involvement–before, during, and after participation–particularly through direct interviews. Addressing this gap may offer valuable insights into the perceived barriers to research participation, inform strategies to enhance population engagement, and promote more diverse representation, which could serve as a foundation for improved recruitment practices that harness the strengths of resilient NICU families.

## Methods

### Recruitment

65 participants from 3 pre-existing NICU research study populations were invited: (1) National Seizure Registry: the Role of Inflammation after neonatal Seizures and later development of Epilepsy (NSR-RISE), (2) The Risk of Acquired Neonatal Significant brain Injury during perinatal Transition in Congenital Heart Disease (TRANSIT-CHD), (3) PRediction Of Maturity, Morbidity, and Mortality in PreTerm infants (PROMPT). The goals of each study are: (1) NSR-RISE (IRB #18-25733) aims to evaluate the relationship between neonatal seizure activity, brain inflammation, and epilepsy development, (2) TRANSIT-CHD (IRB #21-34839) aims to assess differences in brain development in neonates with congenital heart disease, (3) PROMPT (IRB #21-33324) aims to identify metabolic markers to protect preterm infants and predict neonatal mortality and major complications.

Inclusion criteria is:

1. Parent older than 18 years,
2. Child with previous NICU admission,
3. Child was previous participant of NSR-RISE, TRANSIT-CHD, or PROMPT,
4. Child’s age at interview: 18-36 months.

Recruitment occurred via email and phone by the research team and coordinators of NSR-RISE, TRANSIT-CHD, or PROMPT. Of those contacted, 41 participants refused or did not respond. A sample of 24 participants (8 from each study) was achieved. This sample size was determined based on thematic saturation criteria commonly used in qualitative research, through which additional interviews would not yield new thematic insights. Parental data on race, ethnicity, and language was collected from prior studies or during interviews (Table 1). Participants received a $50 gift card.

**Table 1.** Parent participant demographics from NSR-RISE, TRANSIT-CHD, and PROMPT. A total of 24 participants were interviewed.

| Characteristic | NSR-RISE,<br>No. (%) | TRANSIT-CHD,<br>No. (%) | PROMPT,<br>No. (%) |
| --- | --- | --- | --- |
| Sex of Parent: |  |  |  |
| Female | 7 (87.5) | 7 (87.5) | 6 (75.0) |
| Male | 1 (12.5) | 1 (12.5) | 2 (25.0) |
| Race/Ethnicity of Parent: |  |  |  |
| Non-Hispanic/Latino White | 5 (62.5) | 3 (37.5) | 3 (37.5) |
| Hispanic/Latino | 2 (25.0) | 3 (37.5) | 1 (12.5) |
| Black/African American | 0 (0) | 0 (0) | 0 (0) |
| Asian | 0 (0) | 0 (0) | 1 (12.5) |
| Middle Eastern or North African | 0 (0) | 0 (0) | 1 (12.5) |
| Multiethnic | 0 (0) | 1 (12.5) | 2 (25.0) |
| Not Reported | 1 (12.5) | 1 (12.5) | 0 (0) |
| Language Spoken by Parent: |  |  |  |
| English | 7 (87.5) | 7 (87.5) | 8 (100.0) |
| Spanish | 1 (12.5) | 1 (12.5) | 0 (0) |

### Study Design and Data Collection

This qualitative study explored parental perceptions of NICU care and research participation. Semi-structured interviews assessed parents’ backgrounds, NICU/research experiences, and future recruitment recommendations (Figure 1). Survey questions were adapted from Dr. Elizabeth Crouch’s study on NICU parents’ perspectives on autopsy, organ, and tissue donation in the setting of neonatal los.^13^ Interviews were conducted over Zoom (July 2023-August 2024) after obtaining verbal consent. Transcriptions were produced using Zoom’s embedded software, manually reviewed for accuracy, and compared with interview notes. Certified interpreters from University of California, San Francisco were used for Spanish-speaking participants, and all transcripts were anonymized in English. This study was approved by the University of California, San Francisco IRB (#21-33278).

**Figure 1.**
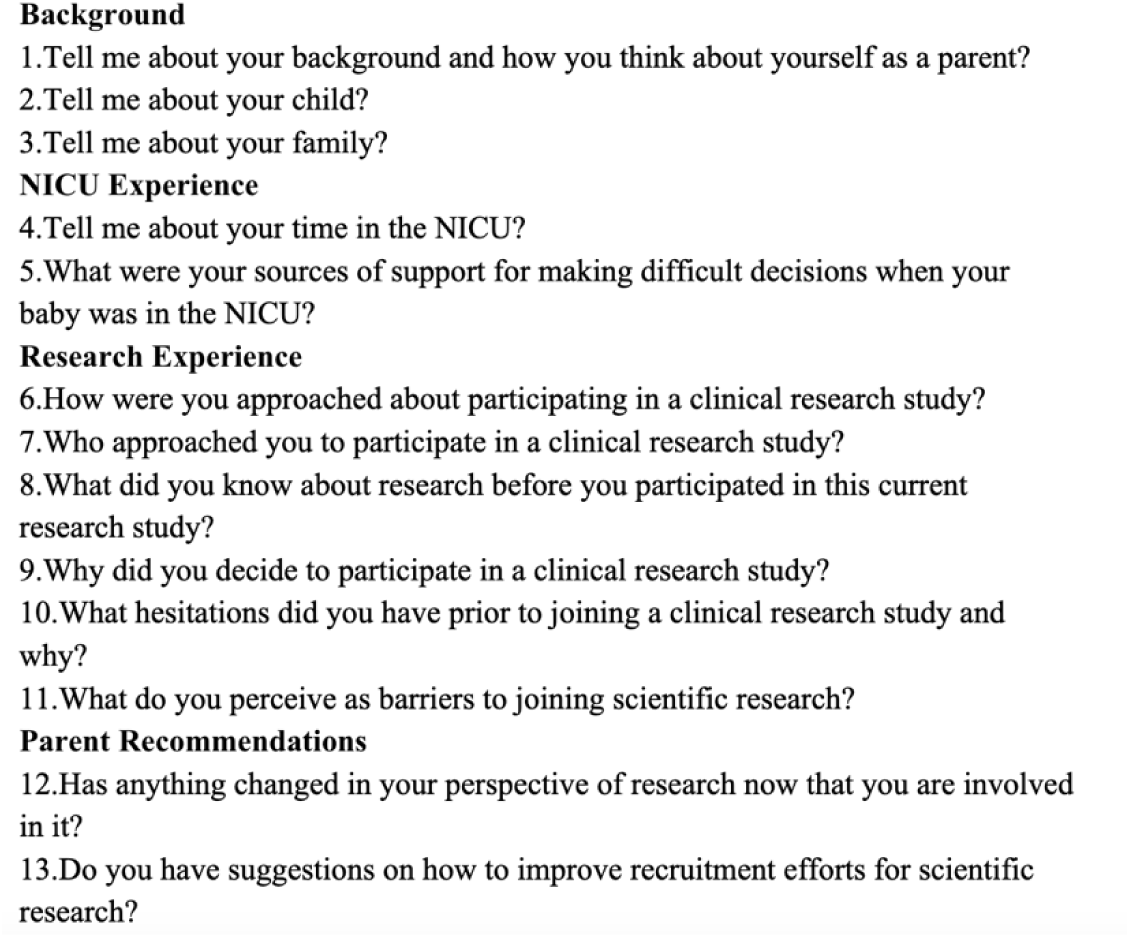
Semi-structured interview questions.

### Data Analysis

A grounded theory approach was utilized, analyzing data prior to hypothesis development to allow themes to emerge organically and explore the NICU parents’ lived experiences without preconceived constraints. Inductive coding identified key codes, sub-themes, and overarching themes directly from the data.

Researchers responsible for coding had advanced training in qualitative analysis, including formal education and completion of specialized training in grounded theory methods. All transcripts were manually coded by multiple authors, with multiple rounds of analysis conducted to ensure reliability. Codebooks modeled after Dedoose software provided a structured coding framework.

Following initial coding, authors met to identify themes, resolving disagreements through discussion until consensus was reached. Thematic saturation, defined as the point when no new themes emerged, marked the conclusion of data collection and analysis. Data analysis occurred from August 2023 to September 2024. Findings were shared with participants and feedback was sought.

## Results

24 participants were interviewed: 8 NSR-RISE parents, 8 TRANSIT-CHD parents, and 8 PROMPT parents, representing 23 different families. Three primary themes emerged: 1) parents’ lived experiences during the emotionally intense NICU period, 2) decision-making regarding NICU research participation, and 3) recommendations for improving NICU research recruitment. An overview of these themes is provided in Figure 3 with exemplary quotes representing these themes shown in Table 2.

**Figure 2.**
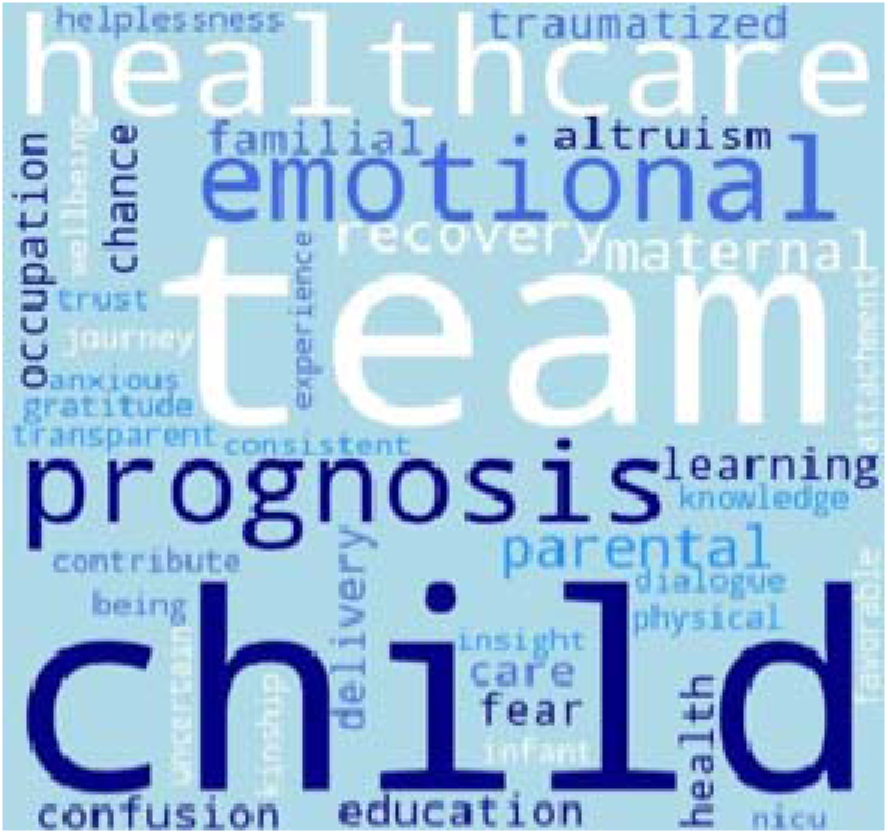
Word cloud.

**Figure 3.**
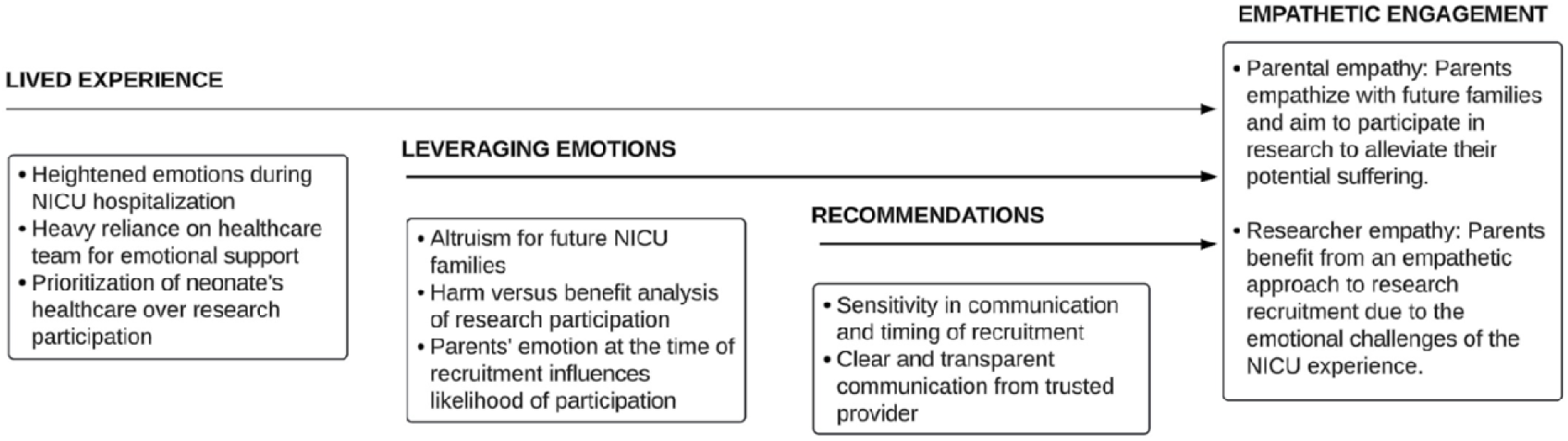
The major themes and subthemes identified in this study emphasize the need for empathetic engagement with NICU families to optimize research recruitment efforts.

**Table 2.** Exemplar quotes.

| Major themes and subthemes | Exemplar Quotes |
| --- | --- |
| **Lived Experience** |  |
| Traumatizing and isolating NICU experience | “Being discharged without a baby...was a different experience...They would tell me ‘You could come see him anytime’ but [I couldn’t]...and it was very unsettling not having him there with me...I felt empty especially after being discharged and carrying this baby with me for 9 months and now I’m leaving the hospital with...empty arms!” |
| Resilience | "[We] can dwell on the fact that it sucks and that it's not fair...and we went through all of those emotions, but at the end of the day...I always bring myself back to the fact that we had the best of the best on our team and how fortunate we were because there's so many people out there that don't have [that]...We felt so well cared for in terms of the knowledge and information that we did receive." |
| Information seeking as primary coping mechanism | “My background of being just a naturally curious person [meant] having information brought security to me...So one experience I had that really molded me...[was] not knowing why things were happening was really scary, and seeing my poor... baby gasping and screaming was a lot for me. I ended up sharing that I wanted to get more information so...one of the residents called me and explained everything to me...All of this didn’t change the outcome...but...just knowing the whole situation, made me feel a lot better...Just learning [and understanding] what's happening...made me feel better, so then I became really active [during rounds] as much as I can, or if there's any studies or anything that I can do, I, I signed up for it." |
| Emotional support | “And I just remember the ICN (NICU) nurse...she just let me cry. And she had...her child in the ICN many years ago, and was just like “This sucks. This sucks and...you just need to feel this, and we're gonna work on it [together].” |
| **Leveraging Emotions** |  |
| Altruism as a key motivator for NICU research participation | “[I know the PROMPT] study won't help [my child], but it'll help other preemies...None of our kids would be here without that research. So why would we not participate? That's my philosophy. So, we said, yes [to participation]...If it's gonna help...someone else's baby in the future...why wouldn't you [participate]? Because you owe your child's life...for everyone that came before. " |
| Increased advocacy | “I [used to just] pull my head down and work and then [my wife] said to me ‘You're not advocating for this!’ ...It's a constant front and sometimes they're a disappointment, sometimes they're successes. So I think this [advocacy] is a new normal more than anything.” |
| Parental understanding of neonatal clinical progress | “I think if [we were recruited to join NSR-RISE] earlier on when we were in the NICU and I didn't know what was going on with my child, I think a barrier would be having the mental and emotional space to even be able to ingest any additional information... I |
|  | remember being in the NICU and the nurse asked if the parent advocate could come talk to me and I almost said no to that because I couldn't imagine talking to anyone else.” |
| **Parent Recommendations** |  |
| Optimizing timing of recruitment | “[I got recruited to join a study while] I was getting a blood transfusion, I was trying to figure out how to pump milk, [my baby] wasn't getting enough breast milk so then we're trying to...decide between formula or donor milk and I was still bleeding a lot...and we still didn't even have the baby because she was still in the NICU! It felt very hard to make any kind of decision in that space, honestly and...I didn't want to say no because of course we want to help but looking back on it...did that have to happen right then?...I do remember feeling like I don't even know where I am right now, never mind can I participate in a study!” |
| Role of trusted provider with established family relationship | “[The researchers] were coming to see him anyway as providers, so they just kind of asked the question after they were done seeing him. As opposed to it being a team of strangers coming and asking for our time... You could maybe have the nurses [that the parents know] ask the parents if maybe they're willing to hear about a study...If it was a complete stranger who we didn't know [recruiting us]...maybe would have not felt that strongly about doing it.” |
| Avoiding medical jargon in communication | “I think it can be kind of intimidating [when you are] not really understanding what exactly, for example, is happening with the blood that they take from [your baby]...There's a lot of big language that's used...like [in] the consent forms...the consent form can be kind of intimidating.” |
| Early communication of study's altruistic goals | “I think that people always want to start with the end game. [If you can start recruitment by saying] ‘If we can figure this out, then we can help the future babies’, who's gonna say no to helping future babies?...And I was never really told what they were trying to do in the future [when I was recruited]...But I think that more people would participate if they knew that there was a chance that they could be helping save lives in the future or enhance the quality of life...And I don't know one person that's gonna be like, ‘No, I don't want that.’” |
| Sensitivity to parental emotional states during recruitment | “[Recruiters should focus on] allowing parents to feel as empowered as possible, because it's a really disempowering experience.” |
|  | “I understand that in intense situations it's harder [for physicians to connect with patients]...But finding those minutes where you can show your humanity, I think, can bring people more closer, and they [parents] can become more open to participating in research.” |

### 1. Parents’ lived experiences during the emotionally intense NICU period

Participants described emotions ranging from gratitude to helplessness during their child’s NICU stay. One parent described, “The [emotions in the] NICU [are] a roller coaster. Some days you’re flying high and some days you’re biting your nails.” Many parents felt isolated during this time, with another parent reporting, “It is the most isolating thing I have ever been through in my life… I backed away from everyone…I felt so alone…That is probably the most…profound and impactful experience that I have been through in my life.” Despite these emotional challenges, many participants demonstrated remarkable resiliency. For instance, one participant remarked, "And now when I think about it, I’m getting emotional…I allow myself to feel [those emotions] and to say, ‘Yeah, that was a lot. That was so much that you were doing [in the NICU]’…and acknowledging, then harnessing it, and then thinking about [the positive] instead of thinking about the worst…Look at him now! He’s brilliant! He’s a smart little guy! He’s chatty and he’s kind. He’s so empathetic. It’s just unbelievable that we have gone through that and we’re here today.” Participants shared their intense emotions occasionally affected their relationships with usual support systems, such as family and friends, leading them to develop new coping strategies, such as information-seeking or building relationships with the NICU team. The NICU team offered education about the child’s diagnosis, prognosis, and treatment options, which many participants found comforting. As one participant noted, “One thing that I personally found really helpful was…I was able to be a part of rounds every day…I liked hearing that conversation…and that helped me not be stressed because I felt like I knew what was happening."

Many participants described a sense of empowerment from their newfound knowledge, leading parents to proactively conduct independent research or consult with individuals with similar experiences. For example, one parent educated herself on her child’s diagnosis and sought guidance from an online support group where she was encouraged to discuss the increased mortality risk associated with delayed treatment with her child’s surgeon. Another participant described advocacy for his family as “a new normal” following their NICU stay. This idea was echoed by another parent, who became an advocate for her child’s ongoing physical therapy needs after discharge.

Participants shared how their parenting styles were transformed after their NICU experience. One particular parent believes her long-term empathy and emotional resonance improved, reflecting “I think there’s a few things that being in the NICU…changed about my approach to her…I give a lot of leeway to her…I don’t know what it’s like to start out life…not being touched by anybody, but it can’t be a great way to start…that’s always in the back of my mind…like what could an experience like that have had on her and what could the result of that be. So, I think I may be more lenient or understanding than I would be of another child.” Overall, while parents’ experiences in the NICU varied significantly, many characterized the experience as profoundly transformative.

### 2. Decision-making regarding NICU research participation

Parents seeking care for their newborns in academic medical centers have the opportunity to enroll their child in research. Participants reported weighing the risks and benefits of enrollment; for example, one parent described, “debating for my child to be a part of a study because of how severe my child may be impacted or in pain and my willingness to say, ‘Please fix my child, even if it’s experimental’ versus ‘I’m not in need of services, my child is happy. There’s no need for me to spend time being a part of a study.’” Participants shared their research participation was influenced by their emotions at the time of recruitment, such as one participant who enrolled her child because of her positivity due to her child’s improving prognosis, stating “I’m not sure if we would have felt the same way about participating in the study if her condition wasn’t progressing well at that point.” This sentiment was echoed by two other participants, who stated, “I think if [recruitment] was earlier on when…I didn’t know what was going on with my child, I think a barrier would be having the mental and emotional space to even be able to ingest any additional information” and “I could see if we were in a different head space, potentially being more hesitant [to participating],” respectively.

Most participants cited altruism as their primary motivator for research participation, expressing deep desire to help future NICU families. One participant enrolled to “see…if my participation could help one more kid in the future… to understand why this happens." Another parent explained, "If it’s gonna help…someone else’s baby in the future…why wouldn’t you [participate]? Because you owe your child’s life…for everyone that came before." One parent emphasized the value of scientific advancement: “The more data that we collect, the more studies that are done on the similar type of issue, the more understanding we have, the more confident we are. And [the more] approaches that we [can] take….”

When asked about barriers to research participation, participants expressed concerns about potential harm to their child. One participant was apprehensive about her child undergoing blood draws, citing “neurodevelopmental outcomes of pain and increased pain being associated with poor outcomes." Many parents reported their child’s NICU study was their first interaction with research, raising additional privacy concerns. Another parent had questions about “her [child’s] personally identified information and what that could…mean for her and some future world.” Financial constraints were also cited, such as one parent whose enrollment was influenced by her husband’s employment situation and lack of sick leave. Other reported barriers included limited medical literacy, language differences, accessibility, and time constraints. Ultimately, NICU parents undergo a complex decision-making process that balances the risks and benefits of study participation.

### 3. Recommendations for improving NICU research recruitment

Participants emphasized the need for better communication and sensitivity to emotional states. While one participant recommended using written materials such as lists, another participant recommended using simpler language and visual aids or diagrams, particularly for individuals with less education: “I’m thinking of one time in particular…a pediatrician…drew out basically the uterus and the baby…to explain to my husband, who did not attend university…so his understanding of the basic sciences is a lot less than me, and that made him become more comfortable.” These suggestions underscore the importance of offering the same information in multiple formats (e.g. lists, diagrams, and illustrations) to accommodate diverse preferences and education levels. Participants also recommended utilizing team members with established relationships with the family for recruitment. One parent appreciated having recruiters who were already involved in her child’s care, stating “If it was a complete stranger who we didn’t know [who recruited us]…maybe [we] would have not felt that strongly about [participating]."

Some participants described instances where recruitment was handled insensitively. One parent described an instance where her child was recruited for a study after difficulty during an MRI: “We had just got him stable, and…[the recruiters] came in like, ‘Oh, it’s simple. We just want to take [his] X-rays.’ And I was like, ‘No. Absolutely not,’…It wasn’t the ask. It was the timing issue on that one, for sure." Similarly, another parent recounted a time when she was approached for recruitment while managing multiple stressors, including a blood transfusion and feeding concerns, stating, “it felt very hard to make any kind of like decision in that space…looking back on it, I’m like ‘Did that have to happen right then?’…I don’t think it was ever explained to me like why that timing had to be right then…I do remember just feeling like I don’t even know where I am right now, never mind can I participate in a study." These experiences highlight that the physical and emotional state of both the parent and newborn impacts how NICU families make their enrollment decisions.

## Discussion

This study explored the complex factors in the NICU parent experience that influence neonatal research enrollment. By examining parents’ perspectives before, during, and after their participation in NICU research, we identified shared features that shape NICU families’ decisions about research participation and areas for improvement. A primary finding was that parents’ decisions to enroll in neonatal research are significantly influenced by their emotional state during recruitment, which is closely linked to the neonate’s clinical status. This underscores the need for empathetic and well-timed communication from trusted researchers and coordination with treatment teams to align recruitment with moments when neonatal prognosis appears more stable.

Effective empathetic communication strategies with NICU parents include providing regular opportunities for questions throughout the consent process, dedicating additional time to hesitant families, and maintaining relationships with families after initial recruitment.^8^ Additional methods include a stepped recruitment approach that uses both verbal and non-verbal cues to gauge parents’ apprehension and adjust the information provided accordingly,^9^ and the use of virtual reality or in-person role-play scenarios.^10^ Alongside empathy training workshops designed for healthcare staff, these tools can cultivate empathetic communication skills.^11^ Our participants prefer when a known team member is responsible for research recruitment, which substantiates Quinn et al.’s finding that leveraging existing relationships between NICU families and healthcare team members can optimize recruitment efforts.^7^ Additionally, Kraft et al. describes building trust with pediatric research participants by determining the optimal time to approach families and remaining adaptable to their needs, ^7^ which our participants also shared as critical to their enrollment decisions. Our study contributes a nuanced understanding of the NICU experience from a parental perspective that is broadly applicable to most NICU parents regardless of the institution they are receiving care, while also highlighting the unique role that research teams play within NICU environments in research institutions. Optimizing the recruitment process not only supports study outcomes but may enhance the overall NICU experience for parents, setting the stage for a collaborative, resourceful family and healthcare network of research studies.

Another key finding from our study is that NICU parents are predominantly motivated by altruism to participate in neonatal research. Nearly every parent interviewed expressed a desire to reduce the challenges faced by future NICU families, aligning with findings from Tromp et al., who identified altruism–specifically the opportunity to aid other families through scientific progress–as a primary motivator for parental research enrollment.^12^ In addition to fostering empathetic communication and carefully timing recruitment efforts, researchers should emphasize altruistic benefits of participation by beginning the recruitment process with the study’s aim to support other NICU families or reinforcing altruistic study goals throughout the consent process. Early identification of potential participating families could also help this process.

Logistical barriers in recruitment can overshadow NICU parents’ willingness to participate in research, perpetuating disparities in neonatal research participation. To enhance diversity and ensure the generalizability of neonatal research, recruitment should actively include a wide range of families, which requires deliberate planning, including having culturally sensitive recruitment staff. Effective and compassionate recruitment in neonatal research should involve a team member who has established rapport or prior contact with the family. This individual should be trained in empathic communication and collaborate closely with the clinical team to identify an appropriate time for recruitment, aligning efforts with the neonate’s clinical progress. Recruiters should employ a variety of communication strategies tailored to parental preferences and education levels, including lists, diagrams, and illustrations, to optimize parental understanding of the study’s goals.

## Limitations

This study is subject to several limitations, such as recall bias due to the 18-to 36-month interval between the newborn’s NICU stay and the parental interview. While this study population included racial diversity and paternal involvement, recruitment of additional non-English speaking participants was limited. Future research should prioritize exploring the impact of cultural differences, including the role of traditional healing practices or cultural perspective on scientific research, on neonatal research recruitment practices and parental preferences.

## Conclusions

NICU patients represent a vulnerable population with variable rates of research participation. Despite the range of complex emotions reported by parents during their child’s NICU hospitalization, many participants expressed willingness to participate in research due to a desire to help other NICU families. To optimize neonatal research recruitment, appropriately timed empathetic communication from trusted persons adapted to parents’ learning preferences, coupled with early emphasis of the study’s altruistic goals, should be prioritized. The intense emotional nature of the NICU experience highlights the need for patient-centered studies that prioritize recruiter flexibility and sensitivity to enhance participant engagement. Aligning recruitment efforts with these motivators may enhance research participation rates and create a network of collaborative families and health professionals who advance research outcomes.

## Data Availability

Data produced in the present study, including codebooks, coding themes, and dialogues are available upon reasonable request from researchers. Please send data requests to authors, Mercedes Paredes or Melissa Coloma

## Acknowledgements

Weston Havens Foundation

The Roberta and Oscar Gregory Endowment in Stroke and Brain Research

The Chan Zuckerberg Biohub

California Preterm Birth Initiative

UCSF Physician Scientist Scholars Program

R01 NS125404 (NIH/NINDS)

